# Lesion distribution and network mapping in dyskinetic cerebral palsy

**DOI:** 10.1101/2024.08.27.24312625

**Authors:** Ana Luísa de Almeida Marcelino, Bassam Al-Fatly, Mehmet S. Tuncer, Ingeborg Krägeloh-Mann, Anne Koy, Andrea A. Kühn

## Abstract

**Objective:** Dyskinetic cerebral palsy (DCP) encompasses a group of predominantly perinatally acquired complex motor disorders that present with dystonia and/or choreoathetosis and are frequently associated with brain lesions in neuroimaging. Recently, lesion network mapping provided a tool to redefine neurological disorders as circuitopathies. In this study, we aim to assess lesion distribution in DCP and identify a DCP-related network derived from lesions.

**Methods:** Here, we review the literature of MRI findings in DCP and perform literature-based lesion network mapping (LNM). Imaging findings and their anatomical distribution were extracted from literature and quantified according to an established MRI classification system for cerebral palsy. Whole-brain functional connectivity from lesions causing DCP was calculated using a pediatric resting-state functional MRI connectome. Results were contrasted with two control datasets for spatial specificity.

**Results:** Review of 48 selected articles revealed that grey matter injury predominated (51%), followed by white matter injury (28%). In 16% of cases MRI was normal. Subcortical lesions affected the thalamus, pallidum and putamen in >40% of reported patients, respectively. Figures available from 23 literature cases were used to calculate DCP-LNM. The LNM revealed functional connectivity to a wide network including the brainstem, cerebellum, basal ganglia, cingulate, and sensorimotor cortices. Strongest connectivity was found for the motor thalamus.

**Interpretation:** The neural network of DCP identified with LNM includes areas previously implicated in hyperkinetic disorders and highlights the motor thalamus as a common network node. The effects of targeting motor thalamic networks with neuromodulation in DCP should be explored in future trials.

## Introduction

Cerebral palsy (CP) is a descriptive term for disorders of movement and posture that result from a non-progressive injury or disturbance to the developing brain.^1^ It can be classified into spastic, dyskinetic and ataxic according to the predominant neurological signs. Dyskinetic cerebral palsy (DCP) represents at least 10-15% of all cases and is characterized by a complex movement disorder including dystonia and/or choreoathetosis.^2–4^ Even though the diagnosis of DCP is not based on etiology, affected children often have a history of hypoxic ischemic encephalopathy (HIE) or, more rarely, bilirubin encephalopathy (BE). Additionally, stroke or cerebral infection can underly the manifestations of DCP in term or preterm born infants.^5^ Currently available treatment options are solely symptomatic and results are variable across patients. Thus, a deeper understanding of the common mechanisms underlying this complex and severely disabling hyperkinetic syndrome represents an urgent unmet clinical need.

Brain imaging shows predominantly grey matter injury including basal ganglia and thalamic lesions.^3,6,7^ However, the exact location within these regions and the frequency of lesions affecting distant areas such as the brainstem, the cerebellum or the hippocampus have not been investigated in detail. Emerging evidence suggests that specific neurological symptoms arise from network malfunction rather than as a consequence of localized brain pathology.^8–10^ For example, manifestation of cervical dystonia in adults has been linked to acquired lesions that were functionally connected to a network comprising the sensorimotor cortex and the cerebellum.^9^ Recently, further networks involved in movement disorders such as parkinsonism and hemichorea have been identified using lesion network mapping.^11,12^ This method assesses whole-brain connectivity from distributed lesions to investigate the network signature of specific neurological and psychiatric symptoms.^8^ In this line, the specific distribution of brain lesions associated with the occurrence of DCP could be harnessed to delineate the neural network that underlies DCP.

Lesion-derived networks were also demonstrated to explain clinical outcome after deep brain sitmulation (DBS) in respective disorders like idiopathic dystonia^9^ or tics^13^. In DCP, the clinical effects of pallidal DBS are variable but overall limited.^14,15^ Patients with more mobile movement components have been suggested to benefit more as compared to those that present with tonic dystonia.^14,16^ Understanding functional networks associated with DCP could pave the way to identify new targets for neuromodulatory therapeutic approaches.

In this study, we sought to use lesion network mapping to trace a functional brain map representative of the DCP network. Furthermore, we considered relevant to provide the reader with a structured overview of the literature of imaging findings in DCP, as it brings our results into the context of current DCP research. Our literature review extends the results reported previously by Aravamuthan *et al.* 2016^6^ by (a) including most recent studies (1990-2023), (b) excluding any restrictions on aetiology of DCP and (c) including only studies assessing imaging through MRI. For lesion network mapping, we used a pediatric resting-state fMRI (rs-fMRI) connectome that has been recently implemented by our research group to enable functional network analyses in pediatric cohorts.^17^

## Materials and Methods

### Systematic Review

A systematic literature review was performed according to the guidelines of the Joanna Briggs Institute for scoping reviews.^18^ The pre-registered review protocol is available online on the Open Science Framework under https://doi.org/10.17605/OSF.IO/N9F8B. Reporting of the scoping review was guided by the adapted PRISMA guidelines (‘Preferred Reporting Items for Systematic reviews and Meta-Analyses’).^19^ Results appear in shortened form in the manuscript (see Figure 1) and can be found in detail in the supplementary files (Supplementary Tables 3, 4).

**Figure 1:**
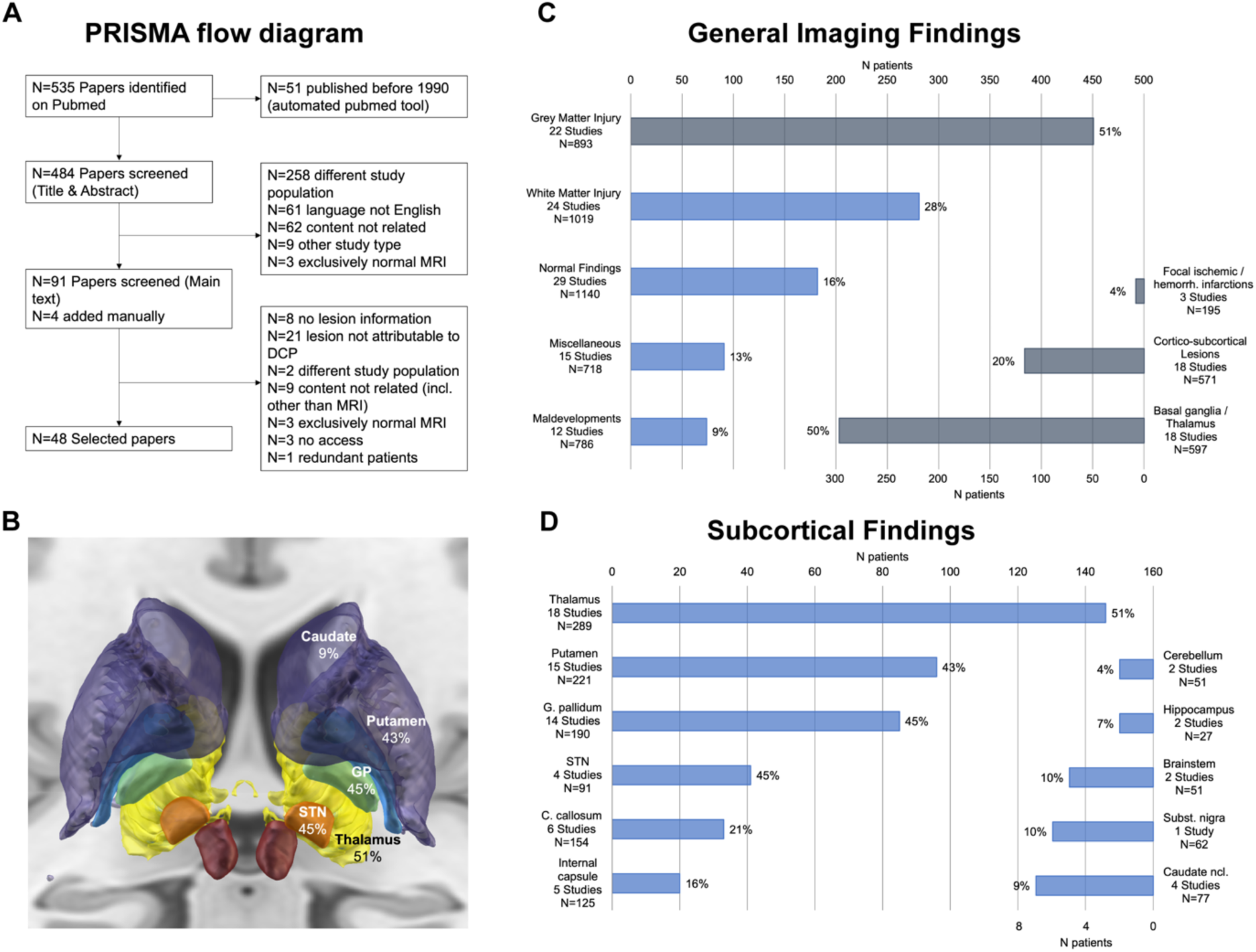
Methods and results of scoping literature review. PRISMA flow diagram for scoping review (A). Anatomical structures with % lesions (B) visualized using DISTAL-Atlas on Lead-DBS.^88,89^ Histogram of lesion distribution in general (C) and in specified subcortical anatomical regions (D).

A literature search was performed in October 2023 in Pubmed (search limitis: 01.01.1990 and 05.10.2023) using “cerebral palsy” and a descriptive term of the typically associated movement disorder such as “dyskinetic”, “dystonia” or “choreoathetosis” as well as “neuroimaging” as search terms (details in Supplementary methods, as in Aravamuthan *et al.* 2016^6^). Full-text peer-reviewed articles written in English that contained MRI findings clearly attributed to this population were included, whereas studies that only included patients with normal MRI findings, where MRI was not performed, not sufficiently reported or not attributable to the DCP population were excluded. Study types considered comprised observational, cohort and case-control studies as well as case series and individual case reports. Experimental/interventional study designs were included when baseline data was available. Reviews, comments, study protocols and editorials were excluded. Titles and abstracts of all studies were first screened and selected by one reviewer (ALAM). The full text of selected citations was reviewed once again for fulfilment of the inclusion criteria and reasons for exclusion were noted. A second reviewer (MT) screened the selected articles and, upon disagreement, discussed it with the first or a third reviewer (BA). References of the selected literature were screened for further suitable articles.

Evidence source data of the selected articles including study date, study type, study population, demographic data, classification system used for lesion description and main outcomes were extracted (Supplementary Table 1). Risk of bias was not specifically assessed. Imaging results, specifically the type of imaging findings and the anatomical localization of lesions, were extracted from the sources of evidence to a customized template adapted from the classification system MRICS^20^, as this is the most widely implemented neuroimaging classification system for cerebral palsy used in large registers.^7,21^ Thus, findings were assigned to the general categories “white matter injury”, “grey matter injury”, “maldevelopments/malformations”, “miscellaneous/not specific” and “normal”. As in MRICS, grey matter injury was further specified, when provided in this form, as “BG/thalamus”, “cortico-subcortical lesions”, “focal ischemic/hemorrhagic infarctions”. Alternatively, the anatomical location of the subcortical injury was described, when given. Thus, if only the specific lesion location was named (e.g. thalamus and putamen), then it would only be reported in the “specific subcortical findings” and not in the “general findings” as grey matter injury. Studies that had one specific lesion pattern as inclusion criteria (e.g. pallidal lesions in Kernicterus in Kitai *et al.*^22^) and have neglected other findings were excluded from the quantitative analysis of imaging findings to avoid reporting bias. Our reporting of the imaging findings included a) number of studies assessing them, b) number of patients included in total in those studies and c) number (and percentage) of patients that have the imaging findings of interest.

### Lesion network mapping

While assessing studies for eligibility, articles were screened for figures of MRI slices which clearly illustrate lesion distribution in children with DCP. We identified 23 cases from 14/48 studies where imaging figures were provided (for a list of the studies included in the connectomic analysis see Supplementary Table 2). Images aquired in infancy (< 1 year of age) or of deformative malformations that could not clearly be traced on MNI space were excluded. All 2D MRI slices extracted from literature were traced manually onto a pediatric MNI template using itk-SNAP (http://www.itksnap.org) and saved as binary lesion masks.^23,24^ Of note, all lesion possible locations belonging to each literature-reported case were traced and saved in a single binary mask even if the lesions were in non-contiguous regions (e.g., putamen and thalamus) or in different slice levels. Functional connectivity seeding from these binary masks to the rest of the brain was estimated for each lesion mask using the Lead-DBS (www.lead-dbs.org) tool Lead Mapper.^25^ Briefly, blood-oxygen-level dependent (BOLD) signal was averaged in the seed region (the binary lesion mask) and correlated to the BOLD signal of each other voxel of the brain. Specifically, the functional connectivity was estimated using a pediatric functional connectome derived from rs-fMRI of 100 neurotypical children^26^ aged 6-18 years implemented in Lead-DBS (https://www.lead-dbs.org/release/download.php?id=PedrsfMRI).^17^ Voxel-wise R values were summarized across the connectome subjects using voxel-wise one-sample t-tests producing voxel-wise T-scores to represent lesion associated connectivity T-maps. Individual T-maps were thresholded at T > 7, binarized and later overlapped to identify a functional network (binary-overlap lesion network map-LNM) common to all lesions associated with the occurrence of DCP.^13^ The T-score thresholding corresponds to an uncorrected *p* < 10^-10^ and a family-wise error-corrected *p* < 10^-4^ in the current pediatric connectome. In order to highlight maximally connected voxels to lesions, the produced LNM was arbitrarily thresholded to demonstrate voxels which have connectivity to at least 95% of lesions. In order to investigate the robustness of the LNM, we calculated another LNM using FSL PALM (https://fsl.fmrib.ox.ac.uk/fsl/fslwiki/PALM). Briefly, a mass-univariate, one-sample t-test across the lesion T-maps was used to derive voxel-wise T-scores harnessing the randomized sign-flipping option in FSL PALM. This statistical analysis is based on permutation testing (5000x) and the voxel-wise resulting p-values were family-wise error (FWE) corrected for multiple comparison with an α<0.05.

### Network-specificity analyses

We compared lesion connectivity profiles (T-maps) of DCP cases to those of two other cohorts. The aim of this comparison is to test the spatial specificity of the previously identified DCP-LNM. First, we used a cohort of focal cortical dysplasia (FCD) that leads to epilepsy in children.^27^ We refer to this cohort as “FCD” cohort. The FCD lesions (n=78) were already traced in native space and made available at (https://openneuro.org/datasets/ds004199/versions/1.0.4). We normalized the anatomical T1w MRI associated with each traced FCD lesion to the pediatric MNI template and applied the warp field to the corresponding tracing. We could only use 77 lesions as a final set in this cohort due to the presence of one lesion outside the brain mask. We then used the normalized tracing as a seed in the pediatric functional connectome and extracted the FCD-lesion associated connectivity profile as above (T-maps). In the second cohort, we created 100 lesion masks in the pediatric MNI template by generating spheres of 12 mm radius and masking them to include only grey matter voxel. We refer to this cohort as “Synth” cohort. We then used the synthesized lesions as seeds in the functional connectome to derive the T-maps. We finally compared DCP-related T-maps to those of the other two cohorts using two-sample t-tests in FSL PALM with 5000x permutations and voxel-wise correction for multiple comparison using FWE with an α<0.05.

## Results

### Selected articles and study characteristics

#### General literature findings

From 535 articles that were initially screened, 91 papers were found eligible for further assessment (reasons for exclusion in Figure 1A). After reading full text, 47 non-suitable articles were excluded and four articles were added manually, yielding a total of 48 articles included in the review. Individual study results and references are provided in the supplementary material (Supplementary Tables 3, 4). Observational studies (23/48) made up the predominant study type and comprised national^28^ and international^7^ register studies. A total of 8318 patients were assessed in all studies, 1901 diagnosed with DCP. Definition of CP subtype was frequently given according to SCPE guidelines (17/48) or not specified (27/48). Most studies included patients with hypoxic-ischemic encephalopathy (HIE), bilirubin encephalopathy (BE) or “other causes” of DCP (36/48 studies). For 12 studies, the cause of DCP was not specified. Age at inclusion ranged from birth to 62 years (mean not calculated since different forms of reporting, available for 44/48 studies), birth weight and gestational age were more seldomly provided (16/48 and 18/48 studies, respectively). Further details regarding study outcomes are summarized in Supplementary table 3.

#### Imaging findings

Age at MRI ranged from day 20 to 62 years (given for 24/48 studies) and reporting of neuroimaging results was standardized using an established classification system such as the MRICS^20^, the preceding classification implemented by Krägeloh-Mann and colleagues^29^ or as in Fiori *et al.*^30^ in 16/48 studies. The first two classifications focus on lesion patterns and presumed timing of the insult, linking the findings to a stage of brain development. The classification by Fiori et. al focuses on neuroanatomical localization and quantification of imaging findings and aims to facilitate structure-function associations, as shown in Laporta-Hoyos et al. 2018^31^ for motor function, communication and cognition in DCP. Reporting of imaging findings was otherwise performed (individually) in extent as in clinical reports^15,32–36^ or guided by generalizable patterns of interest different from the abovementioned established classification systems^29,37–39^. Neuroimaging findings were described to assess the prevalence of specific imaging patterns in CP population studies^3,7,40–43^, to investigate imaging findings in specific etiologies of DCP^22,35,44–51^ or associate them with clinical or functional outcomes^52–55^. In other studies, imaging findings were included to supplement clinical or interventional findings (e.g. DBS, EEG)^2,15,32,34,56–58^ that were the primary focus of the study. Only six studies out of 48 investigated functional^59,60^ or structural^37,38,61,62^ connectivity measures.

### Quantitative analysis and anatomical distribution of imaging findings

A total of eight studies were excluded from the quantitative analysis to avoid reporting bias (details Supplementary Table 4). Thus, data from 40 studies including imaging findings from 1181 DCP patients were available. First, general imaging findings were summarized into the abovementioned categories according to the MRICS classification.^20^ In few cases, exceptions had to be made as two elements of different categories had been summarized together by the authors in the original paper (e.g. “cortical/subcortical” lesions by Préel^63^ include porencephaly, that could be considered a consequence of IVH and would otherwise be assigned to “white matter injury”). An overview of the imaging findings is displayed in Figure 1 (B,C,D). Most common were grey matter injury found in 51% of reported patients (451/893 patients; 22 studies), followed by white matter injury 28% (281/1019; 24 studies), miscellaneous findings 13% (91/718; 15 studies), normal 16% (182/1140; 29 studies) and malformations 9% (74/786; 12 studies). Studies in which grey matter findings could be further specified revealed involvement of basal ganglia with/without thalamus in 50% (296/597; 18 studies), cortical/subcortical lesions 20% (116/571; 18 studies) and focal ischemic/hemorrhagic lesions 4% (8/195; 3 studies).

More than that, 25/40 articles discriminated subcortical findings: here, lesions were mostly found in the thalamus 48% (146/289; 18 studies), pallidum 45% (85/195; 14 studies) and putamen 43% (96/221; 15 studies). Structural abnormalities were also found in further remote areas such as the subthalamic nucleus, the corpus callosum, internal capsule, brainstem, substantia nigra, caudate nucleus, cerebellum, and hippocampus – however, more seldomly reported.

### Network Mapping of DCP-related lesions

Twenty-three cases of patients with DCP with a corresponding MRI figure were identified from 14 articles. The associated lesions are depicted onto a pediatric MNI template in Figure 2. Age at MRI ranged between 1-35 years. The lesions of these 23 cases were heterogeneously spatially distributed in the brain, with a tendency to affect the putamen, the thalamus or both in the majority of cases. Binary-overlap lesion network mapping revealed a functional brain network comprising the brainstem, cerebellum, thalamus, basal ganglia, cingulate and insular cortices as well as the sensorimotor cortex (Figure 3A). This finding was replicated when using one-sample t-test method (Figure 3B) and was also specific to DCP when compared to the FCD and the Synth lesion cohorts (Figure 3C and 3D, respectively). Clusters of thalamic voxels demonstrated the strongest connectivity to lesion location in 95% of DCP cases (Figure 4). The clusters correspond to the anatomical location of the mediodorsal nucleus and the ventral intermediate/ventral oral posterior nuclei (VIM/VOP; Supplementary Figure 4). We sought to further segregate the functional part of the thalamus to which the lesions were maximally connected. As such, a post-hoc analysis exploring the connectivity strength of each lesion to the four functional thalamic parcellations (namely, motor, limbic, associative and other brain regions) was conducted (see Supplementary Methods). Since there is no pediatric functional parcellation of the human thalamus available, we built a thalamic parcellation atlas from the 100 subject normative pediatric connectome. A group comparison with analysis of variance (ANOVA) revealed a significant effect of functional connectivity to the different thalamic parcellations (p = 0.0002). Post-hoc analyses via paired t-tests showed that functional connectivity to the sensorimotor parcellation of the thalamus was significantly higher than to associative and limbic parcellations (p < 0.001 in both cases; Bonferroni correction; see Figure 4B, right, for illustration and reported p-values). Since some of the cases present thalamic lesions, we controlled for redundancy by repeating the analysis “masking out” the thalamic lesions as seeds. This control analysis revealed a similar thalamic cluster to that of the original analysis (Supplementary Methods and Supplementary Figure 2).

**Figure 2:**
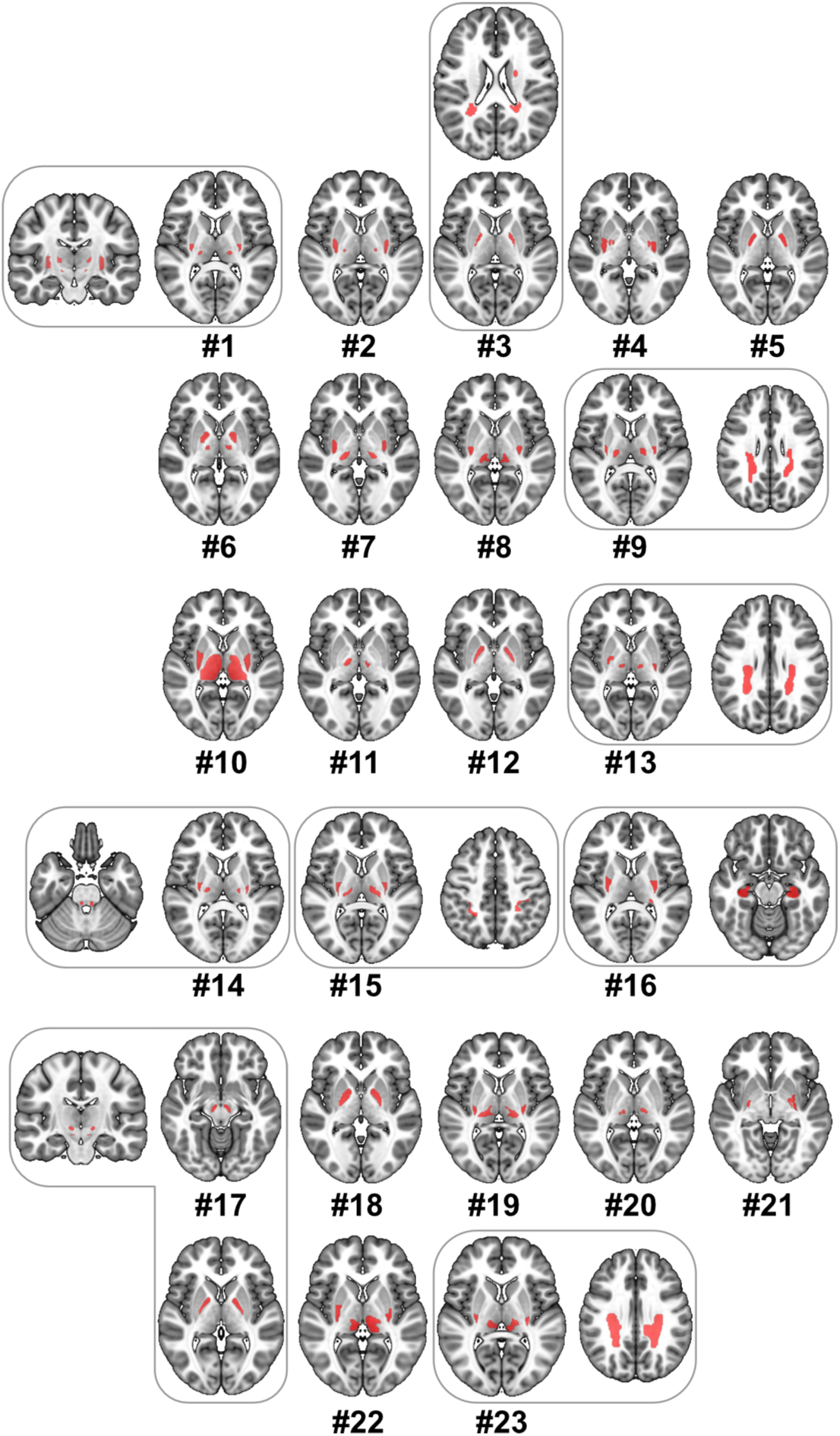
Spatial distribution of lesions associated with DCP. Each #Number represents an individual case from literature. Lesions were heterogenously distributed in the putamen, globus pallidi, thalamus and white matter. Case #1 and #17 have additional subthalamic nucleus lesions, while case #14 has a brainstem lesion. It is noteworthy to mention that combined lesions occurring in single cases were taken as a single seed in the connectomic analysis.

**Figure 3:**
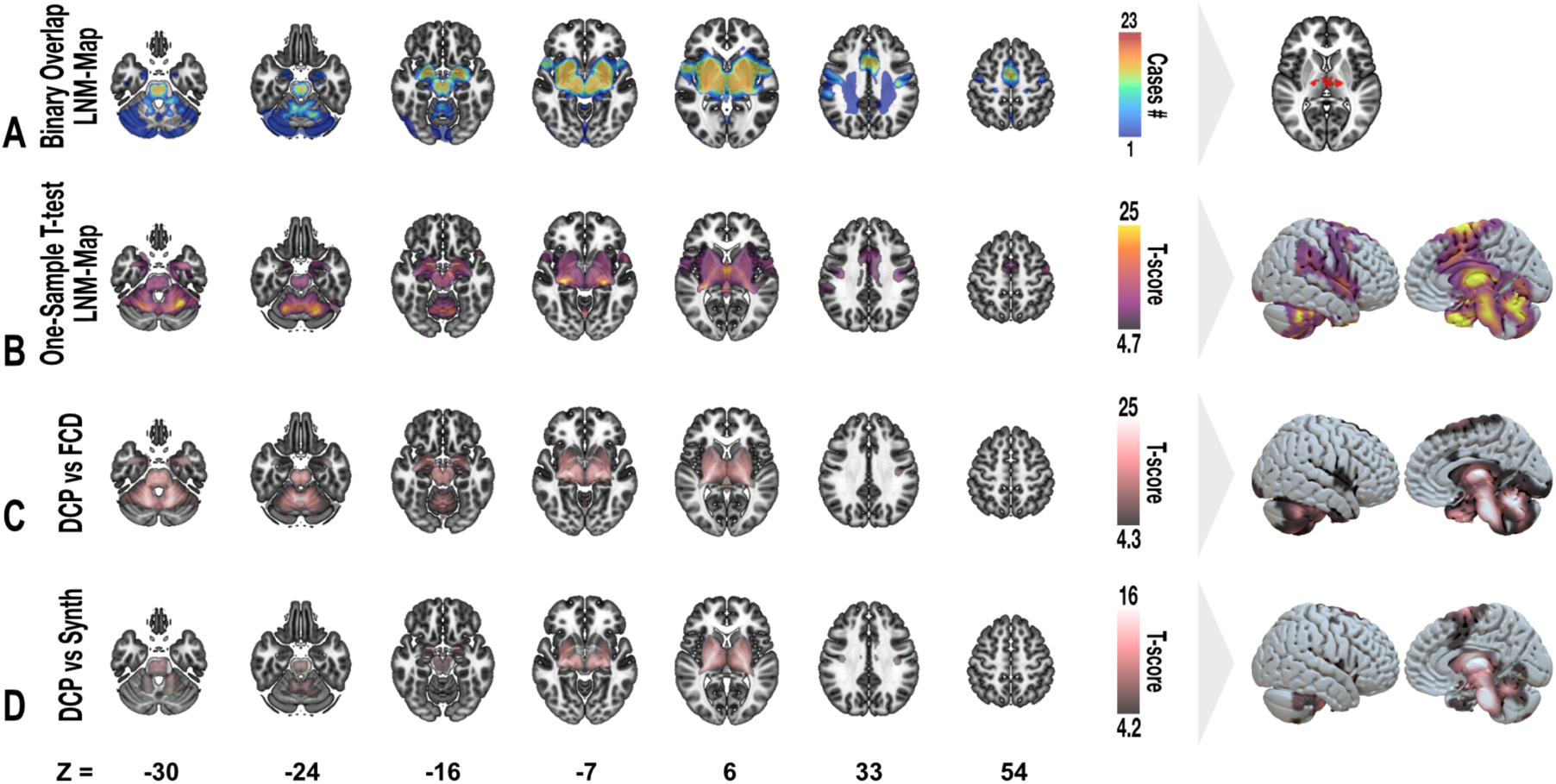
Lesion network mapping of DCP. Different mapping methods have been used to replicate the resulting LNM for DCP. A binary-overlap of T-maps thresholded to T > 7 is depicted in (A) which is later strictly thresholded to show only voxels of high connectivity to lesions. By running a permutation-based, one-sample t-test over all DCP-lesion T-maps, a network of significant voxels was determined which is statistically robust (B). Comparing DCP-lesion associated connectivity profiles to that of focal cortical dysplasia (C) and a synthesized set of lesions (D) emphasized that DCP-LNM is also specific to DCP. All voxel-wise t-scores (B-D) were corrected for multiple comparison using family-wise error using a *p*-value of < 0.05. Backdrop of the volumetric slices is the skull-stripped T1w MRI modality of the pediatric MNI (4.5-18.5yr) template.^23^ Surface rendering of the maps is depicted using a surface mesh of pediatric MNI template.

**Figure 4:**
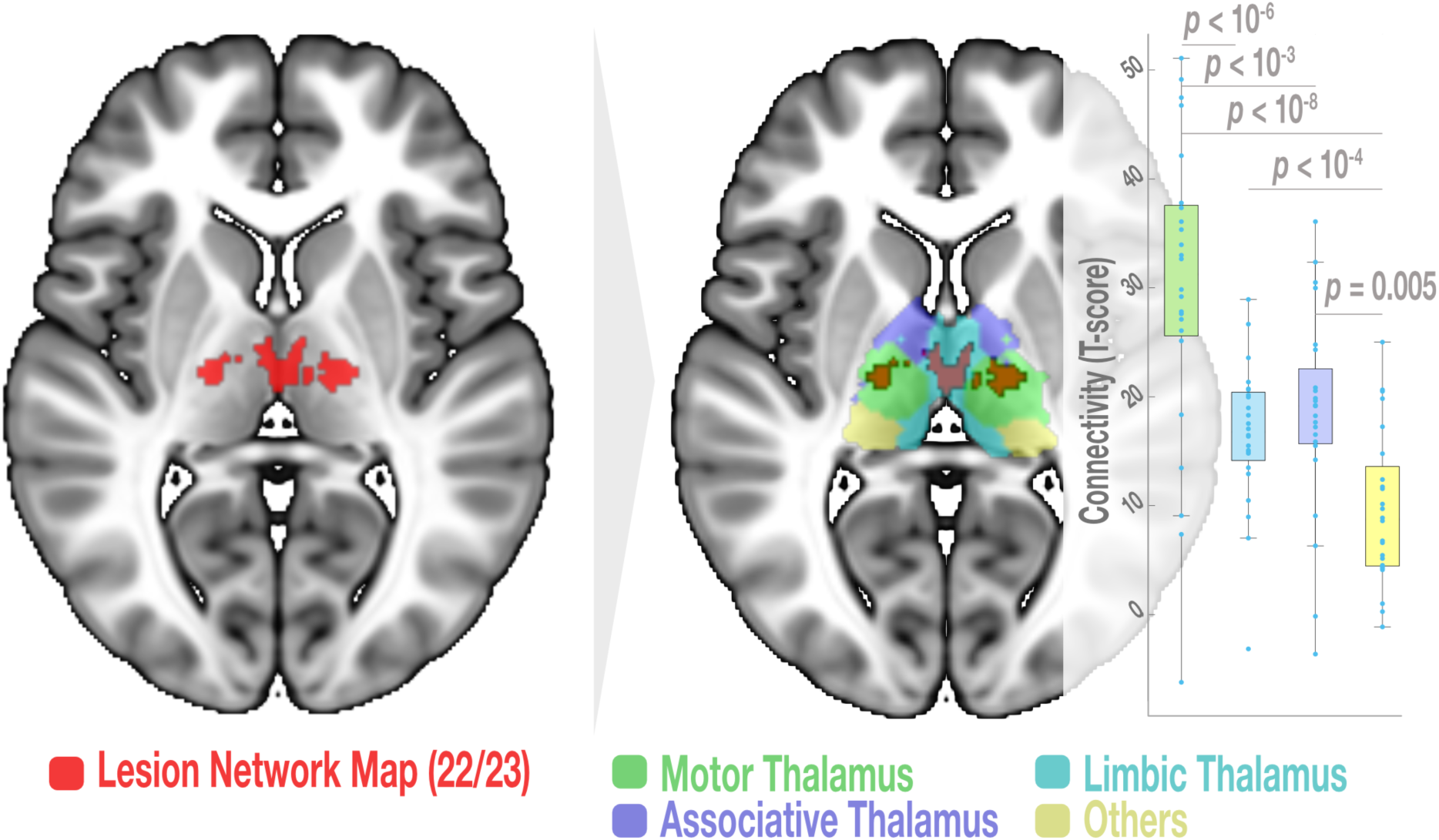
The thalamus as an important node of DCP-associated LNM. Mapping lesions associated with DCP indicated a cluster of highest connectivity (connected to 22/23 cases) located in the thalamus (left panel in red). This cluster was overlaid on a “winner-takes all” functional thalamic parcellation (right panel) extracted using predefined cortical regions of interest (motor, associative, limbic and other brain regions) in the 100 subjects of the pediatric rs-fMRI connectome. Boxplots represent the connectivity between each single lesion and each of the thalamic parcellation. Connectivity to the motor thalamus was significantly higher than to any of the other thalamic parcels. The pairwise t-test *p*-values were reported as Family-wise error-Bonferroni corrected *p*-values.

## Discussion

In this study, we performed a structured, comprehensive review of lesion distribution in DCP to delineate a lesion network map associated with this complex movement disorder. We show that while evidence of imaging findings in DCP comes predominantly from observational studies assessing the prevalence of general imaging patterns, specific lesion distribution is less frequently reported and includes remote subcortical areas. Our results of LNM reveal a distributed network including common motor network nodes such as the basal ganglia, cerebellum and sensorimotor cortex that have been previously implicated in hyperkinetic movement disorders. Importantly, the LNM reported here converges in the motor thalamus, which extends our current understanding of this disabling disease.

### Clinicoanatomic insights from brain lesions in DCP

DCP is clinically defined by its presentation with hyperkinetic motor symptoms dominated by dystonia and choreoathethosis.^1,4^ The underlying etiology is variable as well as the lesions that have been described in patients diagnosed with DCP. Reviewing the literature, we show that most often the prevalence of specific imaging patterns is assessed in relation to cerebral palsy subtypes^7,41,64,65^ (spastic or dyskinetic), different etiologies^22^ or gestational ages^7,63^ in observational (population-based) studies. Commonly used imaging classification systems underline the association between injury patterns and stages of cerebral development that has been proposed in the past.^20,66^ Our review results corroborate previous studies that reveal grey matter injury to be the most common imaging finding in DCP.^7^ An increased vulnerability of putaminal and thalamic neurons to hypoxia in the perinatal period as observed in HIE^67,68^ or of pallidal neurons to neurotoxic bilirubin as in BE^69^ can explain frequent structural damage of these regions in DCP. Furthermore, individual differences in neuronal vulnerability or variability in the exposure to the damaging event may lead to different lesion patterns within the same DCP etiology.^70,71^ In contrast to classical periventricular white matter injury that mostly disrupts the corticospinal tract and leads to spasticity, the basal ganglia are part of a more intricate circuit with complex local and remote connections, thus leading to more complex phenotypes.^72^ For example, it is known that basal ganglia and thalamic injury patterns due to inherited (metabolic) diseases cause similar complex dyskinetic phenotypes as seen in DCP.^73^ Also, mutations in genes involved at different hubs of the basal-ganglia-thalamocortical loop can lead to complex dyskinetic movement disorders and mimic cerebral palsy through cell-specific effects that do not necessarily reflect tangible lesion patterns in the MRI.^74^

In this study, we focus on the common clinical presentation of DCP as a complex dyskinetic movement disorder and deliberately set no restrictions on its etiology. Evidence regarding the association between specific motor phenotypes in DCP and imaging findings is scarce. While mixed type DCP (dyskinesia and spasticity) has been associated with isolated or concomitant white matter injury, patients with isolated dystonia frequently present isolated basal ganglia lesions.^53^ Monbaliu and colleagues^2^ observed a higher incidence of choreoathetosis in patients with “pure” thalamus and basal ganglia lesions in comparison with mixed lesions (e.g. additional white matter lesions). However, specific movement disorder patterns or dyskinesia severity have not been systematically associated with specific imaging findings, for instance using the dyskinesia impairment scale or kinematic analyses.

Our literature review shows that lesions leading to the clinical manifestation of DCP can be heterogenously distributed within the basal ganglia nuclei or thalamus but can also affect remote locations such as the corpus callosum, internal capsule, brainstem, hippocampus or cerebellum. Within the framework of movement dosorders being considered network disorders we postulate that despite different putative lesion mechanisms and lesion distribution they have in common the perturbation of a DCP-specific functional network.

### Towards a network signature of DCP

Recent advances in brain mapping and connectomic analyses have contributed to redefine neurological diseases as network disorders. Lesion network mapping (LNM) is exemplifying this paradigm shift in the approach to neurological diseases and has allowed to identify relevant brain networks in different conditions including movement disorders.^9,10,13^ The current study is a further use case of LNM in a pediatric neurological disorder. Our results partially corroborate previous findings of LNM in chorea and dystonia in adult populations.^9,12^ Regions like the cerebellum, anterior cingulate cortex, thalamus, globus pallidus, putamen and the brainstem were indeed parts of the LNM in chorea and dystonia. Although the motor cortex was negatively correlated in terms of connectivity to lesions causing dystonia, it was positively connected to lesions causing chorea in adults.^9,12^ Noteworthy, even though terminology of childhood movement disorders is derived from adults, specific features in clinical presentation differ and underlying mechanisms cannot be transferred one-to-one.^4^ The aforementioned regions comprise important hubs in motor control previously associated with complex hyperkinetic syndromes of different etiologies in childhood.^74^ Perturbation of such networks during a critical timepoint in brain development adds complexity to the symptoms encountered in children as compared to focal lesions to the adult brain.^75,76^ The use of a pediatric connectome in our study aimed to account for this relevant aspect in pediatric disorders.

Previous studies have demonstrated evidence of structural or functional changes in DCP patients in different regions of the LNM demonstrated here. Qin and colleagues found reduced interhemispheric connectivity in motor network areas including the cerebellum, motor cortex, supplementary motor area and anterior cingulate cortex^60^ as well as altered functional connectivity within cerebellar, sensorimotor and left frontoparietal resting state networks^59^ in patients with DCP. Structural connectivity studies indicated reduced white matter integrity in motor tracts including the corticospinal tract as well as cerebellar connections and association fibres related to speech and language.^37,38^ On a structural network level, reduction of white matter connectivity measures was associated with impaired motor and cognitive skills.^62^ Altered functional and structural connectivity measures indicate a more widespread impact of brain lesions on neural networks that are not always captured by conventional MRI measures. Importantly, the current LNM included areas showing hypometabolism in DCP caused by HIE and Kernicterus, namely the cerebellum, the brainstem, the putamen and the thalamus (Supplementary Figure 4 depicts significant peak regions).^77,78^ This convergent evidence implies an important pathophysiological contribution of these regions to DCP and indicates involvement of a widespread neural network beyond the structurally damaged areas. This finding is also relevant to cases of DCP without imaging abnormalities (as in 16% of cases identified in our review process), for which one could speculate on a subtle metabolic lesion or dysfunction on a cell-specific level that affects the here deciphered LNM even if no structural damage is detectable on imaging.

The finding that almost all DCP lesions included in the current study are highly connected to the motor thalamus marks the central role of this particular hub of the motor network. In fact, thalamic stroke has been previously shown to cause complex hyperkinetic movement disorders^79^. The thalamus integrates information from the basal ganglia, the cerebellum and sensory afferents in complex circuitries to inform cortical dynamics.^80^ Even though traditionally assigned to the basal ganglia, encoding of action selection and movement vigor in the motor thalamus has been shown to influence movement kinematics in rodents.^81^ Also, the temporal dynamics of thalamo-cortical interactions are crucial for intact motor execution.^82^ Thus, altered spatio-temporal precision of signal processing throughout the identified network may be one possible explanation for manifestation of complex hyperkinetic movement disorders after lesions structurally or functionally affecting the motor thalamus and could be tested in electrophysiological studies.

### Implications for future studies

The results of this study could have relevant implications for advancing DCP diagnosis and treatment. Following our approach to DCP as a motor network disorder independent of its etiology, future studies can further elucidate the impact of specific lesion patterns on brain networks to eventually predict patients at risk and symptom severity in DCP. In addition, applying connectomics to larger DCP cohorts could allow disentangling different motor phenotypes and respective imaging findings. This, along with knowledge regarding treatment responses may help to better understand the underlying pathophysiology of DCP and allow identification of new targets for neuromodulatory interventions such as DBS. Currently, the globus pallidus pars internus is commonly targeted for DBS in childhood dystonia of different etiologies, including DCP.^83^ However, due to variable clinical effects^14,15^ and anatomical complexity associated with the presence of lesions, alternative targets such as the VIM/VOP thalamic nuclei^84^ and recently the dentate nucleus^58^ have been suggested. Intriguingly, all these DBS targets lie within the DCP-lesion network identified in our study (Supplementary Figure 4 depicts the relationship of DCP-LNM to alternative DBS targets). Of note, DCP causative lesions demonstrated the highest connectivity to the VIM/VOP target. Even though the pallidum and the dentate display different connectivity strengths, they reside within the identified DCP network. The association of lesion-derived and therapeutic networks has been demonstrated for tics and dystonia in adults with Tourette syndrome^13^ and isolated idiopathic dystonia^9^, respectively. In DCP, the implanted DBS system should counteract the dysfunctional network effect of lesions. Lastly, the motor cortex is another hub of the LNM identified here which could serve as a target for non-invasive stimulation techniques. In one study, cathodal transcranial direct current stimulation over the motor cortex was probed to downregulate excessive activity found in this area with variable success in controlling motor symptoms in a small sample size.^85^ Assessing the impact of such therapeutic options would require further prospective clinical trials.

### Limitations

Literature-based studies are limited by the fact that they attempt to summarize a heterogeneous body of evidence that has emerged due to different questions and using different approaches. Reporting of imaging findings was standardized using previously established classification systems in >30% of studies included, yielding higher comparability between studies. However, focusing on predominating lesion patterns leads to a reporting bias, thus possibly neglecting important structural findings.^20,30^ The definition of (D)CP has not been stated in all studies which could affect generalizability of results. There is a consensus that diagnosis of CP should only be made after the second year of life, however we did not make this restriction in this study. Furthermore, advances in diagnostic methods such as exome sequencing have yielded different diagnoses in up to 30% of CP patients.^86^ These data refer, however, to convenience samples, which were not population based and in part highly selected. In addition, lesion location at a “macro” scale only explains part of variance in clinical manifestation. HIE, but also genetic syndromes that cause specific lesion patterns (e.g. PDE10A mutations) and can be included in DCP cohorts, present a specific neuronal vulnerability at a “micro” scale that will not be sufficiently reflected in imaging patterns. Lastly, performing LNM from literature is limited to 2D figures. However, it has been demonstrated that seeding from 2D lesions in functional connectomes is similar to 3D volumetric lesions.^87^ Also, the use of a normative connectome does not fully reflect the network effects lesions would have on the individual brain they occur on.

## Conclusion

This study characterizes a widespread DCP functional lesion network highlighting the motor thalamus as a main network node. The DCP neural network overlaps with previously proposed pathophysiological concepts in hyperkinetic conditions and provides additional insights into potential therapeutic targets. Future work should focus on testing the utility of the identified network in predicting clinical outcome and therapeutic benefits in prospective DCP cohorts.

## Acknowledgements

A.L.A.M., B.A. and A.A.K. are supported by Collaborative Research Centre TRR 295 (Project ID 4247788381) of the “Deutsche Forschungsgemeinschaft” (DFG). A.A.K. is supported by DFG under Germanýs Excellence Strategy EXC-2049 – 390688087 and additionally funded by the Lundbeck Foundation as part of the collaborative project grant “Adaptive and precise targeting of cortex-basal ganglia circuits in Parkinsońs Disease” (Grant Nr. R336-2020-1035). A.K. is funded by the University hospital of Cologne. She holds grants from the medical faculty and the Dr. Hans-Günther and Dr. Rita Herfort foundation. M.S.T. is supported by the Edmond J. Safra Fellowhip for Movement Disorders of the Michael J. Fox Foundation. The funding sources had no involvement in study design, in the collection, analysis, and interpretation of data; in the writing of the manuscript, and in the decision to submit the paper for publication.

## Author contributions

A.L.A.M, B.A. and A.A.K. contributed to the conception and design of the study. A.L.A.M., B.A., M.T. and A.A.K. contributed to the acquisition and analysis of data. A.L.A.M., B.A., M.T., I.K.M. and A.K. contributed to drafting the text and preparing the figures.

## Data availability

Lesion tracings and lesion network map are available from the corresponding author upon reasonable request. All code used is available within Lead-DBS/-Connectome software (https://github.com/netstim/leaddbs).

## Supplementary material

Supplementary material is available *online*.

