## Supplementary for "Lesion distribution and network mapping in dyskinetic cerebral palsy"

#### Supplementary Methods

**Searching Terms for Scoping Review**

a. ((cerebral palsy) OR (acquired dystonia))
b. AND (dyskinesia OR dyskinetic OR chorea OR choreiform OR dystonia OR dystonic OR athetosis OR athetotic OR athetoid OR choreoathetosis OR choreoathetoic OR choreoathetoid)
c. AND (MRI OR imaging OR neuroimaging OR lesion)
d. NOT (Review[Publication Type])

**Functional parcellation of the thalamus**

Since the cluster identified in lesion network mapping of dyskinetic cerebral palsy was located inside the thalamus, a post-hoc analysis to determine which functional thalamic subregion was highly connected to the lesions was necessary. To do so, we used the 100 subjects from the normative resting-state functional pediatric connectome [1] to parcellate the thalamus in the into four main functional zones, namely the motor, associated, limbic and other brain regions. First, *cortical* masks (or regions of interest) representative of the motor, associative, limbic and other brain regions were built by combining different cortical regions from the Haskins pediatric atlas [2]. No subcortical and cerebellar regions were included in the region of interest masks. In order to do so, we warped the Haskins brain template and its associated atlas to the pediatric MNI space and later selected single cortical regions to be contributing to each cortical mask. Specifically, the postcentral paracentral, and precentral gyri were merged to form the motor cortex mask [3]. The limbic cortex mask was extracted based on merging of the orbitofrontal, frontopolar cortices and the anterior cingulate gyri, while the associative cortex comprised all the remaining prefrontal cortices. All other brain regions (other parietal, temporal and occipital regions) were merged into one functional category as “others”. Of note, Haskins to pediatric MNI spatial warping was carried out by linearly co-registering (using SPM[4]) and nonlinearly normalizing (using ANTs, http://stnava.github.io/ANTs/[5]) the Haskins template to the pediatric MNI template in Lead-DBS toolbox[3]. The resulting warp field was then applied to the associated Haskins annotated atlas. We then run functional connectivity from the combined cortical masks (motor, associative, limbic and others) to the rest of the brain in each of the pediatric normative connectome subjects. This yielded 100 connectivity maps from each cortical mask (in total 4 x 100 maps). The maps were then masked by a thalamic binary image extracted from the pediatric Haskins atlas. Voxels were assigned as belonging to a specific parcel using a winner takes all strategy[6]. The winner takes all method ensures that a specific thalamic voxel is highly connected to the specific cortical region across the 100 subjects of the pediatric normative connectome. The distribution of the thalamic parcels was visualized and compared to that of published adult thalamic parcellation by overlaying the final parcellation masks onto a pediatric MNI template[7] (Supplementary Figure 1). The spatial distribution of the parcels conforms to a similar spatial distribution when compared to its adult counterpart[8] (https://github.com/BrainMappingLab/Functional-territories-of-basal-ganglia). Note that the adult parcellation is based on structural imaging data.

### Supplementary Figures

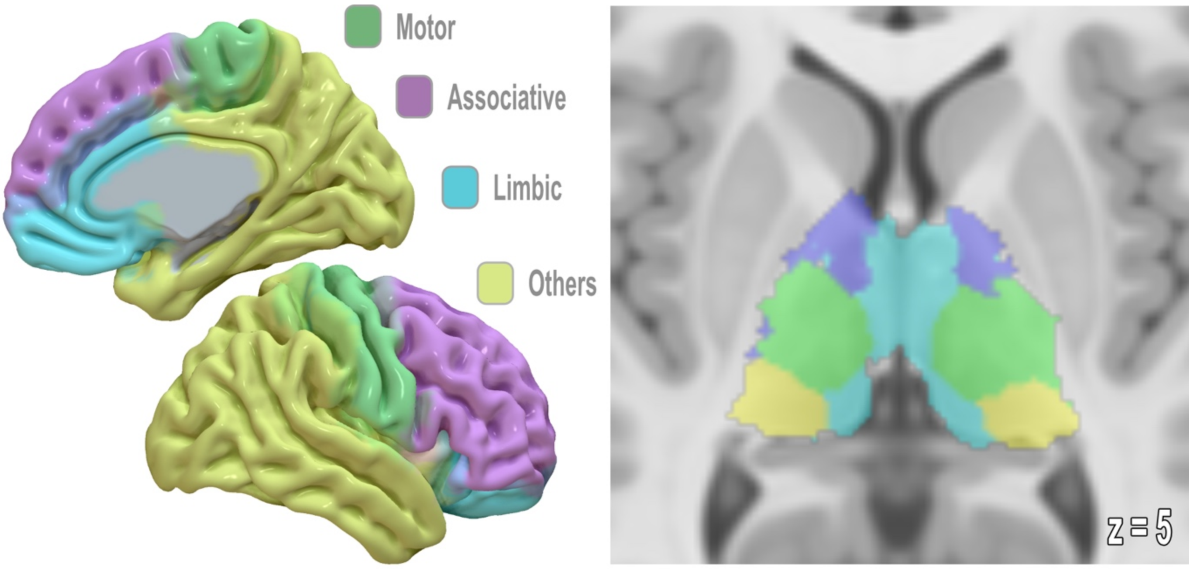

**Supplementary Figure 1: Functional Parcellations of the Pediatric Thalamus.** Cortical functional zones (left panel) were formed by merging multiple brain regions into one category. Motor (green), associative (purple), limbic (cyan) and others (yellow) masks where then used as seed regions in subject-wise connectivity mapping using the rs-fMRI acquisitions from the 100 neurotypical subjects in the pediatric connectome. The resulting maps were masked to extract only information inside thalamic voxels and later grouped into four different thalamic parcels using winner takes all parcellation scheme (right panel).

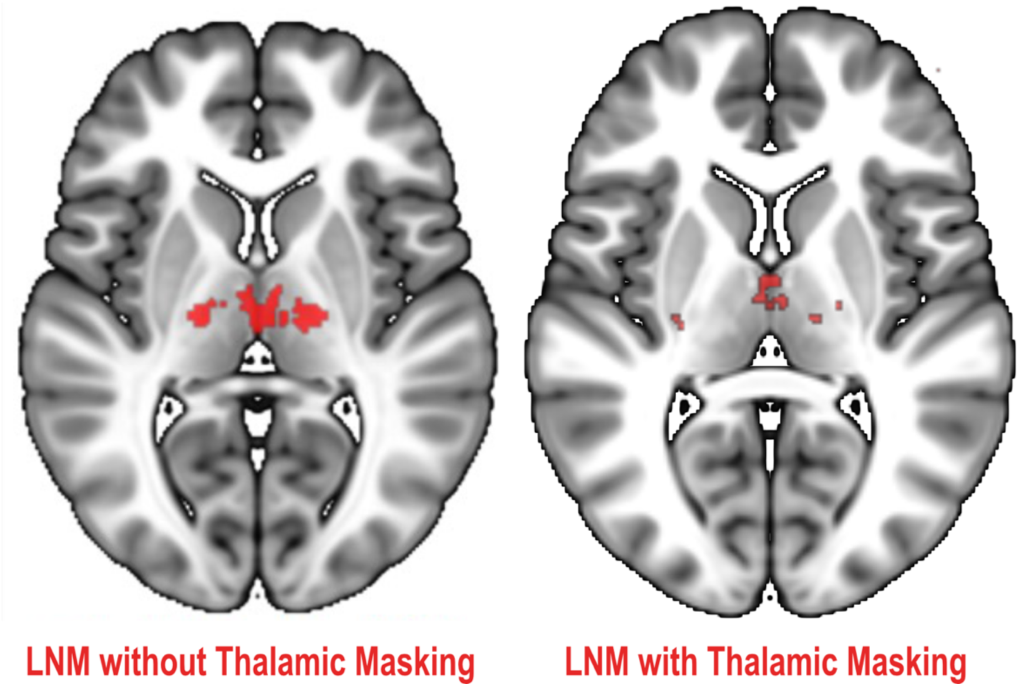

**Supplementary Figure 2: LNM Control Analysis.** Masking out thalamic lesions did not differently affect LNM spatial distribution.

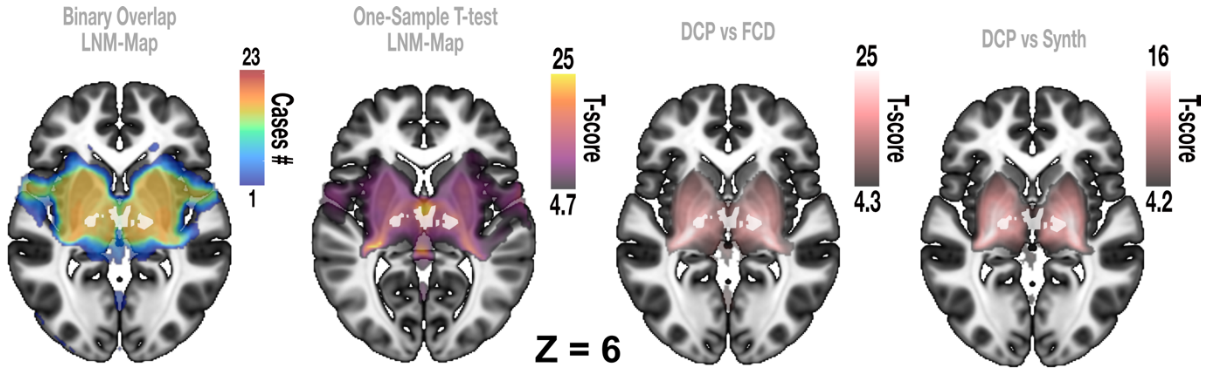

**Supplementary Figure 3: Specificity analysis.** Overlap of the highest connectivity cluster (white) on different DCP-related maps. This cluster lies within both comparison (specificity) maps (last two panels) when using FCD or synthesized lesions. The cluster can also be replicated using the One-sample t-test LNM-mapping method (second panel).

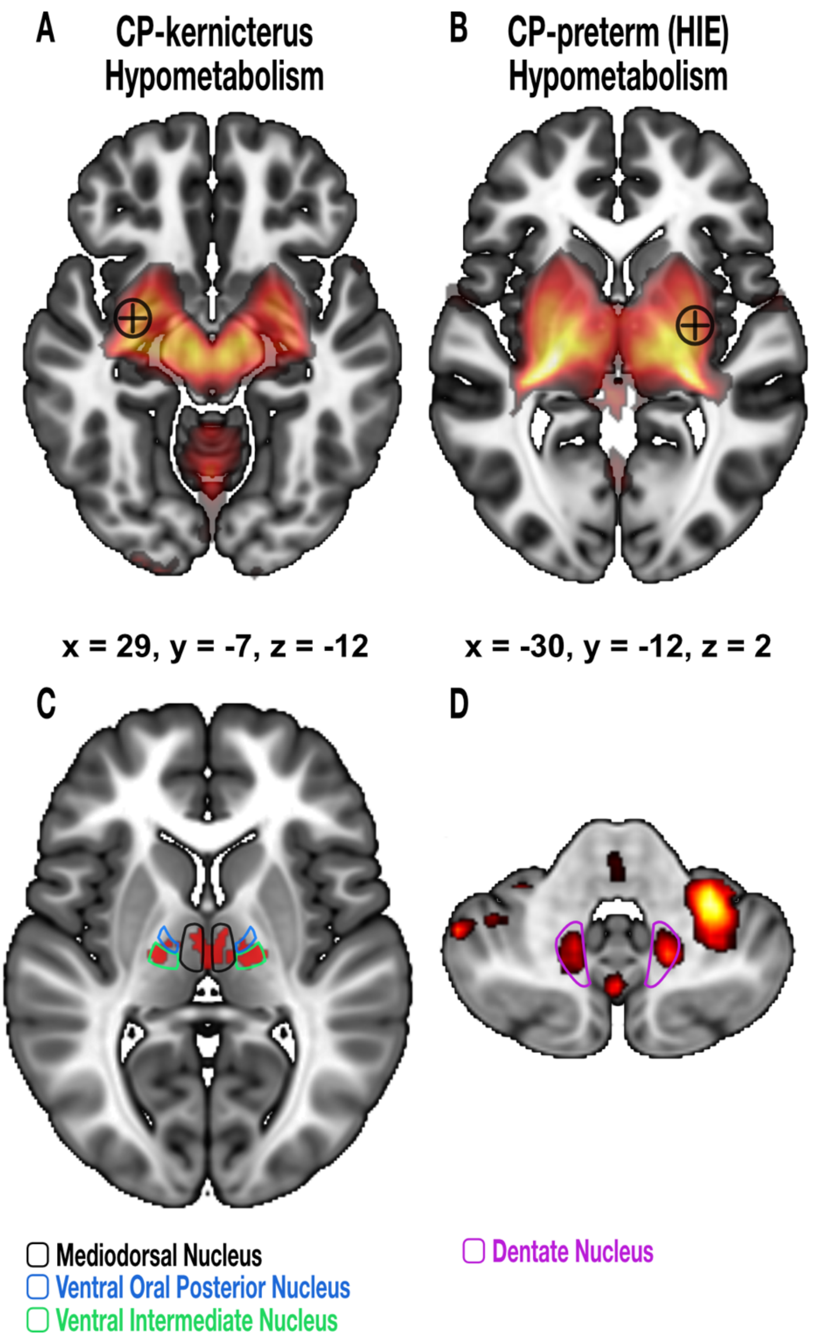

**Supplementary Figure 4:** Relationship of one-sample t-test LNM comparison map to hypometabolism coordinates identified by Tsagkaris and colleagues[9] using FDG-PET (A and B). Both coordinates of peak hypometabolism occurring in CP-kernicterus and hypoxic ischemic encephalopathy (HIE) cases were depicted with a black circle and are mentioned below each axial slice. The coordinates are taken from the supplementary material of Tsagkaris et al.[9] The thalamic clusters highly connected to DCP lesions were located in the expected spatial location of the mediodorsal nucleus for the medial cluster and the ventral intermediate and ventral oral posterior nuclei for the lateral cluster (C). The lateral cluster agrees with a deep brain stimulation target to treat dystonia in children including DCP. The dentate nucleus, which is a newly used target for deep brain stimulation in DCP is also overlapping with clusters identified in the one-sample t-test LNM (D). The latter LNM is highly thresholded to a T-score of (13/20) to better show the overlap.

### Supplementary Tables

**Supplementary Table 1:** Data extraction table for scoping review.

| |
|---|
| Study Title |
| First Author |
| DOI |
| Year of Publication |
| Study Type |
| Study Population |
| Number All Included Patients |
| Number Patients DCP |
| Aetiology of CP included (Hypoxic Event/ Hyperbilirubinaemia/ Other) |
| Age at inclusion |
| GA (weeks) |
| Birth Weight (g) |
| Imaging Modality (Field+Sequence) |
| Number of DCP with MRI |
| Images Provided (y/n) |
| Lesions assigned to DCP |
| Systematic Lesion Classification |
| Lesions Report (Table/Text) |
| Outcome Structural Imaging |
| Outcome CP Subtype |
| Outcome Classification Scale (GMFCs, MACS, CFCS) |
| Outcome Motor Scale (DIS, BFMDRS,…) |
| Outcome Other (structural/functional/intervention) |
| Video reported |

**Supplementary Table 2:** Summary of the 23 cases of DCP identified from literature which have traceable imaging slices.

| Paper | Cases | Figure | age | sex | Clinical presentation | Lesion etiology |
|---|---|---|---|---|---|---|
| **Saini et al. 2021** | | | | | | |
| | Case #1 | 1 | // | // | Dyskinetic CP *(not further specified)* | Hyperbilirubinemia |
| | Case #2 | 2 | // | // | Dyskinetic CP *(not further specified)* | Perinatal asphyxia |
| **Kitai et al. 2021** | | | | | | |
| | Case #3 | 2A | // | // | Dyskinetic CP *(not further specified)* | Bilirubin encephalopathy |
| | Case #4 | 2B | // | // | Dyskinetic CP *(not further specified)* | Hypoxic-ischemic encephalopathy |
| **Park et al. 2013** | | | | | | |
| | Case #5 | 2B | 33 | male | GMFCS: 4 BADS: 18 | // |
| | Case #6 | 2C | 23 | female | GMFCS: 2 BADS: 19 | // |
| | Case #7 | 2D | 35 | male | GMFCS: 4 BADS: 22 | // |
| | Case #8 | 2E (1-2) | 16 | female | GMFCS: 2 BADS: 19 | // |
| **Yokochi et al. 1991** | | | | | | |
| | Case #9 | 1A-B | 6 | female | Athetotic CP | // |
| | Case #10 | 2 | 3 | female | Athetotic CP | // |

| | | | | | | |
|---|---|---|---|---|---|---|
| | Case #11 | 3 | 3 | male | Athetotic CP | // |
| **Sun et al. 2018** | | | | | | |
| | Case #12 | 1 (upper) | 1 | female | Dystonic CP | Hyperbilirubinemia |
| | Case #13 | 2 (mid) | 8 | male | Choreoathetotic CP | Hypoxic-ischemic encephalopathy |
| **Derinkuyu et al. 2016** | | | | | | |
| | Case #14 | 2 (a-d) | 3 | female | Dyskinetic CP *(not further specified)* | Hypoxic-ischemic encephalopathy |
| **Hoyos et al. 2018** | | | | | | |
| | Case #15 | 3 | 14 | male | Dyskinetic CP *(not further specified)* | // |
| **Griffith et al. 2009** | | | | | | |
| | Case #16 | 3 | // | // | Dyskinetic CP *(not further specified)* | Hypoxic-ischemic encephalopathy |
| **Koy et al. 2014** | | | | | | |
| | Case #17 | 1 | 22 | // | General dystonia + spasticity<br>GMFCS: 5<br>BFMDRS-M: 100<br>BFMDRS-D: 26 | Peripartal asphyxia |
| **Gkoltsiou et al. 2008** | | | | | | |
| | Case #18 | | 1 | female | Mild dyskinetic CP | Hyperbilirubinemia (G6PD-deficiency) |
| **Cajigas et al. 2023** | | | | | | |
| | Case #19 | 1A | 19 | female | Dyskinetic CP<br>BFMDRS:<br>51 (DBS OFF)<br>31 (DBS ON) | Perinatal hypoxia |
| **Okumura et al. 2006** | | | | | | |

| | | | | | | |
|---|---|---|---|---|---|---|
| | Case #10 | 1 | 4 | male | Athetoid CP | Kernicterus |
| **Yilmaz et al. 2002** | | | | | | |
| | Case #21 | 1 | 1 (16 months) | male | Dyskinetic cerebral palsy and limitation of vertical gaze | Kernicterus |
| **Menkes et al. 1994** | | | | | | |
| | Case #22 | 2 | 5 | // | Dyskinetic CP, mental retardation | Hypoxic-ischemic encephalopathy |
| | Case #23 | 3 (A-D) | 6 | // | Dyskinetic CP (not further specified) | Perinatal asphyxia |

**Supplementary Table 3:** Characteristics of studies included in the scoping review. Background blue for general literature characteristics, green for demographics of included patients, orange for imaging-related results and grey for non-imaging outcomes. Reference of studies with specific result given (according to Suppl. Table 4).

| ***Study Characteristics*** | **Results of Scoping Review** | **Nr. Studies (%)** |
|---|---|---|
| *Publication date* | 1991-2023 | All |
| *Study type* | Observational (cross-sectional/populational) [1,3,5-8,10,11,15,18-21,26-31,33,35,41,48] | 23 (48) |
| | Retrospective (not further assignable) [2,4,9,12,14,17,23,25,32,34,38] | 11 (23) |
| | Prospective non-interventional [13,16,22,24] | 4 (8) |
| | Case series [36,37,40,42,46] | 5 (10) |
| | Case reports [39,43,44,47] | 4 (8) |
| | Randomized controlled trial [45] | 1 (2) |
| *Total patients included* | 8318 | - |
| *Total DCP included* | 1901 | - |
| *Study population* | Primarily patients with DCP [3,5-9,12,16-18,21-23,27,29,33,36,37,40,43-47] | 24 (50) |
| | ≥ 2 CP subtypes [1,2,4,10,11,13,15,19,20,25,28,30-32,35,38,48] | 17 (35) |
| | Other (e.g. “childhood dystonia” or “children at risk for CP”) [14,24,26,34,39,41,42] | 7 (15) |
| *Age at inclusion* | Birth to 62 years | 44 (92) |
| *Gestational age* | 23-41 weeks | 18 (38) |
| *Birth weight* | 620-4500g | 16 (33) |
| *Cause of CP* | HIE, BE and other [1,3,7,10,14,16-19,21,24,27,30,31,33,35] | 16 (33) |
| | HIE only [26,32,36,39,41,45-47] | 8 (17) |
| | BE only [12,23,34,40,42-44] | 7 (15) |
| | HIE and BE [5,9] | 2 (4) |
| | HIE and “other” [28,37,38] | 3 (6) |

| | | |
|---|---|---|
| | Cause not specified [2,4,6,8,11,13,15,20,22,25,29,48] | 12 (25) |
| *CP-subtype definition* | According to SCPE guidelines [1-3,6-8,10,11,13,16,18,20-22,29-31,35] | 17 (35) |
| | According to other definitions [11,38,46,48] | 4 (8) |
| | Not specified | 27 (56) |
| *Imaging modality* | Conventional MRI | 42 (86) |
| | - Structural connectivity measures [2,4,8,16] | 4 (8) |
| | - Functional connectivity measures (rs-fMRI) [13,22] | 2 (4) |
| | MRI and [CT and/or ultrasound] [6,18,30,31,35,48] | 6 (13) |
| *Age at MRI* | Day 20 – 62 years | 24 (50) |
| *Reporting of imaging results* | Systematic established classification (e.g. MRICS) [1,3,6,8,11,18-20,27,29-32,35,38,45] | 16 (33) |
| | Other (e.g. in extent, using unpublished patterns of interest) | 32 (67) |
| *Non-imaging outcomes* | Functional classification scales (GMFCS, MACS, CFCS, other) | 23 (48) |
| | - Only GMFCS [1-3,8,10,13,15,22,24,41,45,48] | 12 (25) |
| | - GMFCS and [MACS and/or CFCS and/or other] [5-7,11,12,16,18,20,21,27,29] | 11 (23) |
| | Quantification of motor symptoms using scores/scales | 17 (35) |
| | - "Burke Fahn Marsden Dystonia Rating Scale" (BFMDRS) [7,9,14,24,33,37,45] | 7 (15) |
| | - "Barry-Albright Dystonia Scale" (BADS) [3,8,14,18] | 4 (8) |
| | - "Dyskinesia Impairment Scale" (DIS) [6,7,26,45] | 4 (8) |
| | - "Alberta Infant Motor Scale" (AIMS) [33,41] | 2 (4) |
| | Prevalence of CP subtype (dyskinetic/spastic/ataxic) in population [1,10,11,15,19,20,25,28,30,31,35,48] | 12 (25) |
| | Non-motor outcomes (quality of life, activities of daily living, cognition) [9-11,16,21,22,27,29,33,34,38,41] | 12 (25) |
| | Electrophysiological measures (motor evoked potentials, EMG, EEG) [7,8,24] | 3 (6) |
| | Therapeutic intervention (DBS) [9,14,34,39,47] | 5 (10) |

**Supplementary Table 4:** Title, year of publication, first author and DOI of all papers included in the scoping review. Articles highlighted in grey were excluded from quantification of imaging findings since a) exact number of DCP patients receiving MRI was not reported (n=3)[1,11,15] or b) they only assessed a specific imaging feature, and it is not clear if other findings were absent or just not reported (n=5)[5,12,25,38,40].

| Number | Title | Year of Publication | First Author | DOI |
|---|---|---|---|---|
| 1 | Which is the Most Common Physiologic Type of Cerebral Palsy? | 2022 | Kamate | https://doi.org/10.4103/0028-3886.349640 |
| 2 | Anatomical characterization of athetotic and spastic cerebral palsy using an atlas-based analysis | 2013 | Yoshida | https://doi.org/10.1002/jmri.23931 |
| 3 | Hyperbilirubinemia and Asphyxia in Children With Dyskinetic Cerebral Palsy | 2021 | Saini | https://doi.org/10.1016/j.pediatrneurol.2021.02.002 |
| 4 | Athetotic and spastic cerebral palsy: anatomic characterization based on diffusion-tensor imaging | 2011 | Yoshida | https://doi.org/10.1148/radiol.11101783 |
| 5 | Functional outcomes of children with dyskinetic cerebral palsy depend on etiology and gestational age | 2021 | Kitai | https://doi.org/10.1016/j.ejpn.2020.11.002 |
| 6 | Clinical patterns of dystonia and choreoathetosis in participants with dyskinetic cerebral palsy | 2015 | Monbaliu | https://doi.org/10.1111/dmcn.12846 |
| 7 | Using both electromyography and movement disorder assessment improved the classification of children with dyskinetic cerebral palsy | 2022 | Lorentzen | https://doi.org/10.1111/apa.16152 |
| 8 | Neuroradiological and neurophysiological characteristics of patients with dyskinetic cerebral palsy | 2014 | Park | https://doi.org/10.5535/arm.2014.38.2.189 |
| 9 | Pallidal stimulation for acquired dystonia due to cerebral palsy: beyond 5 years | 2014 | Romito | https://doi.org/10.1111/ene.12596 |
| 10 | Children with dyskinetic cerebral palsy are severely affected as compared to bilateral spastic cerebral palsy | 2019 | Préel | https://doi.org/10.1111/apa.14806 |
| 11 | Dyskinetic vs Spastic Cerebral Palsy: A Cross-sectional Study Comparing Functional Profiles, Comorbidities, and Brain Imaging Patterns | 2018 | Reid | https://doi.org/10.1177/0883073818776175 |
| 12 | Diagnosis of Bilirubin Encephalopathy in Preterm Infants with Dyskinetic Cerebral Palsy | 2020 | Kitai | https://doi.org/10.1159/000502777 |

| | | | | |
|---|---|---|---|---|
| 13 | Functional Connectivity Alterations in Children with Spastic and Dyskinetic Cerebral Palsy | 2018 | Quin | https://doi.org/10.1155/2018/7058953 |
| 14 | Pallidal stimulation in children: comparison between cerebral palsy and DYT1 dystonia | 2013 | Marks | https://doi.org/10.1177/0883073813488674 |
| 15 | Clinical and MRI correlates of cerebral palsy: the European Cerebral Palsy Study | 2006 | Bax | https://doi.org/10.1001/jama.296.13.1602 |
| 16 | Whole-brain structural connectivity in dyskinetic cerebral palsy and its association with motor and cognitive function | 2017 | Ballester-Plané | https://doi.org/10.1002/hbm.23686 |
| 17 | Magnetic resonance imaging in athetotic cerebral palsied children | 1991 | Yokochi | https://doi.org/10.1111/j.1651-2227.1991.tb11955.x |
| 18 | Dyskinetic cerebral palsy: a population-based study of children born between 1991 and 1998 | 2007 | Himmelmann | https://doi.org/10.1111/j.1469-8749.2007.00246.x |
| 19 | Magnetic resonance imaging, risk factors and co-morbidities in children with cerebral palsy | 2010 | Prasad | https://doi.org/10.1007/s00415-010-5782-2 |
| 20 | The Origin of the Cerebral Palsies: Contribution of Population-Based Neuroimaging Data | 2020 | Horber | https://doi.org/10.1055/s-0039-3402007 |
| 21 | Clinical characteristics and functional status of children with different subtypes of dyskinetic cerebral palsy | 2018 | Sun | https://doi.org/10.1097/md.0000000000010817 |
| 22 | Aberrant Interhemispheric Functional Organization in Children with Dyskinetic Cerebral Palsy | 2019 | Qin | https://doi.org/10.1155/2019/4362539 |
| 23 | Kernicterus in preterm infants | 2009 | Okumura | https://doi.org/10.1542/peds.2008-2791 |
| 24 | EEG measures of sensorimotor processing and their development are abnormal in children with isolated dystonia and dystonic cerebral palsy | 2021 | McClelland | https://doi.org/10.1016/j.nicl.2021.102569 |
| 25 | A magnetic resonance imaging finding in children with cerebral palsy: Symmetrical central tegmental tract hyperintensity | 2017 | Derinkuyu | https://doi.org/10.1016/j.braindev.2016.10.004 |
| 26 | Dyskinesia Impairment Scale scores in Dutch pre-school children after neonatal therapeutic hypothermia | 2020 | Kuiper | https://doi.org/10.1016/j.ejpn.2020.07.013 |
| 27 | Brain lesion scores obtained using a simple semi-quantitative scale from MR imaging are associated with motor function, communication and cognition in dyskinetic cerebral palsy | 2018 | Laporta-Hoyos | https://doi.org/10.1016/j.nicl.2018.06.015 |

| | | | | |
|---|---|---|---|---|
| 28 | Regional cerebral glucose metabolism in clinical subtypes of cerebral palsy | 1991 | Kerrigan | https://doi.org/10.1016/0887-8994(91)90024-f |
| 29 | Prevalence, birth, and clinical characteristics of dyskinetic cerebral palsy compared with spastic cerebral palsy subtypes: A Norwegian register-based study | 2023 | Evensen | https://doi.org/10.1111/dmcn.15598 |
| 30 | The panorama of cerebral palsy in Sweden part XII shows that patterns changed in the birth years 2007-2010 | 2018 | Himmelmann | https://doi.org/10.1111/apa.14147 |
| 31 | The changing panorama of cerebral palsy in Sweden. X. Prevalence and origin in the birth-year period 1999-2002 | 2010 | Himmelmann | https://doi.org/10.1111/j.1651-2227.2010.01819.x |
| 32 | Anatomic localization of dyskinesia in children with "profound" perinatal hypoxic-ischemic injury | 2010 | Griffiths | https://doi.org/10.3174/ajnr.a1854 |
| 33 | Young adults with dyskinetic cerebral palsy improve subjectively on pallidal stimulation, but not in formal dystonia, gait, speech and swallowing testing | 2014 | Koy | https://doi.org/10.1159/000360984 |
| 34 | Serial brain MRI and ultrasound findings: relation to gestational age, bilirubin level, neonatal neurologic status and neurodevelopmental outcome in infants at risk of kernicterus | 2009 | Gkoltsiou | https://doi.org/10.1016/j.earlhumdev.2008.09.008 |
| 35 | The panorama of cerebral palsy in Sweden part XIII shows declining prevalence in birth-years 2011-2014 | 2022 | Himmelmann | https://doi.org/10.1111/apa.16548 |
| 36 | Athetoid cerebral palsy with cysts in the putamen after hypoxic-ischaemic encephalopathy | 1992 | Rutherford | https://doi.org/10.1136/adc.67.7_spec_no.846 |
| 37 | Cerebellar deep brain stimulation for the treatment of movement disorders in cerebral palsy | 2023 | Cajigas | https://doi.org/10.3171/2023.1.JNS222289 |
| 38 | Bilateral lesions of thalamus and basal ganglia: origin and outcome | 2002 | Krägeloh-Mann | https://doi.org/10.1017/s0012162201002389 |
| 39 | Brain magnetic resonance imaging in suspected extrapyramidal cerebral palsy: observations in distinguishing genetic-metabolic from acquired causes | 1997 | Hoon | https://doi.org/10.1016/s0022-3476(97)70160-4 |
| 40 | Single photon emission computed tomography and serial MRI in preterm infants with kernicterus | 2006 | Okumura | https://doi.org/10.1016/j.braindev.2005.11.004 |
| 41 | Motor testing at 1 year improves the prediction of motor and mental outcome at 2 years after perinatal hypoxic–ischaemicencephalopathy | 2009 | van Schie | https://doi.org/10.1111/j.1469-8749.2009.03302.x |

| | | | | |
|---|---|---|---|---|
| 42 | Magnetic resonance imaging in three children with kernicterus | 2001 | Sugama | https://doi.org/10.1016/S0887-8994(01)00306-X |
| 43 | Thalamic involvement in a patient with kernicterus | 2002 | Yilmaz | https://doi.org/10.1007/s003300100993 |
| 44 | Proton magnetic resonance spectroscopic images in preterm infants with bilirubin encephalopathy | 2012 | Kamei | https://doi.org/10.1016/j.jpeds.2011.09.036 |
| 45 | Quality of Life After Deep Brain Stimulation of Pediatric Patients with Dyskinetic Cerebral Palsy: A Prospective, Single-Arm, Multicenter Study with a Subsequent Randomized Double-Blind Crossover (STIM-CP) | 2023 | Koy | https://doi.org/10.1002/mds.28898 |
| 46 | Clinical and MR correlates in children with extrapyramidal cerebral palsy. | 1994 | Menkes | PMID: 8197940 (DOI not available) |
| 47 | Bilateral Thalamic Lesions in a Newborn with Intrauterine Asphyxia After Maternal Cardiac Arrest — a Case Report with Literature Review | 2001 | Banerjea | https://doi.org/10.1038/sj.jp.7210560 |
| 48 | Population-based study of neuroimaging findings in children with cerebral palsy | 2011 | Towsley | https://doi.org/10.1016/j.ejpn.2010.07.005 |

### References (Supplementary Material)
